# The role of plasmapheresis in snake envenoming: a systematic review

**DOI:** 10.1101/2024.09.27.24314476

**Authors:** Hari Prasad, Nidhi Kaeley, Jewel Rani Jose, Ajun U N, Takshak Shankar, Ajmal Salam, Krishna Shukla

## Abstract

**Background:** Envenoming from numerous sources, such as snakes, scorpions, and spiders, is a major health issue across the world, resulting in millions of cases and tens of thousands of deaths annually. Venom induced symptoms ranges from systemic reactions like nausea and vomiting to localised pain and swelling. One major risk is the development of venom induced consumption coagulopathy (VICC), which might result in significant consequences. Plasmapheresis is being investigated as a possible therapy for severe envenoming.

**Objectives:** We aim to assess the effectiveness and potential advantages of plasmapheresis in snakebite cases, focusing on clinical results. We seek to find if plasmapheresis improves neurological, renal, and hematological dysfunction and impacts secondary outcomes, including patient discharge rates, morbidity, mortality, duration of hospital stay, and the number of plasmapheresis sessions required.

**Methods:** Following PRISMA guidelines, we conducted a systematic search of articles published between 1980 and July 2023 across multiple databases. MeSH terms related to snakebite and plasmapheresis were applied without publication or language type restrictions. Inclusion criteria considered case reports, cross-sectional studies, or case series featuring plasmapheresis in snakebite management. Inclusions were participants aged 18 years or older with confirmed or suspected snakebites, meeting plasmapheresis indications. Exclusions included participants under 18 years, studies reporting only in vitro data, review articles, and redundant reporting. The emphasis was on Emergency Departments or Intensive Care Units.

**Results:** In a review of 147 cases (1980 to July 2023), the most common snake was the hump-nosed viper (*Hypnale hypnale)*. Renal, neurological, and hematological dysfunctions improved after plasmapheresis. The mean plasmapheresis sessions were 2.1, and the average hospital stay was 13.13 days.

**Conclusion:** Once the data has been analyzed, the result emphasizes the clinical importance of plasmapheresis in snakebite envenoming. It helps decision-making when standard therapies are insufficient or ineffective, potentially saving lives.

**Author Summary:** Snakebites pose a significant global health threat, causing numerous deaths and serious injuries annually. While antivenom is the primary treatment, it’s not always effective or available. This study explores an alternative treatment called plasmapheresis, a method that filters harmful substances from the blood.

We reviewed 147 cases of snake envenoming treated with plasmapheresis between 1980 and 2023. Our findings show that plasmapheresis can improve various complications caused by snake venom, including kidney problems, nerve damage, and blood disorders. On average, patients received about two plasmapheresis treatments and stayed in the hospital for around 13 days.

The study suggests that plasmapheresis could be a valuable option when standard treatments aren’t working well enough. It might help save lives in severe cases of snake envenoming. While more research is needed, this review provides important insights for doctors treating snakebite victims, especially in areas where snakebites are common and resources are limited.

## Background

Envenoming causes significant morbidity and mortality; the most common causes are snakes, scorpions, spiders, bugs, and unidentified bites [1]. According to the World Health Organisation (WHO), there are 2.7 million snake bites globally each year, resulting in 81,000 to 138,000 deaths [2]. Snakebite is classified as a category A Neglected Tropical Disease (NTD) by the WHO [3]. Each year, there are over 81,000 occurrences of snakebite envenoming in India, resulting in 11,000 fatalities [4].

Indian Cobra (Naja Naja), Russell’s Viper (*Daboia russelii)*, and Krait (*Bungarus caeruleus)* are among the most harmful and venomous snakes. Examples of local symptoms include painful regional lymphadenopathy, edema, and discomfort at the bite site. Nausea, vomiting, headaches, diaphoresis, stomach discomfort, and diarrhoea are examples of non-specific systemic symptoms. Toxin syndromes such as neurotoxicity, myotoxicity, and VICC can result from systemic envenoming [5]. Some elapid envenomings cause hemostatic and cardiovascular consequences [6].

Acute kidney injury (AKI), VICC, and tissue destruction are all caused by metalloproteinases, phospholipase A2, and snake lectins, in hematotoxic venom. The major toxin-mediated coagulopathy brought on by a hemotoxic snakebite is VICC, which is then followed by venom-induced Thrombotic Microangiopathy (TMA) and Diffuse Alveolar Hemorrhage (DAH) [7,8]. Russell’s viper (*Daboia russelii)* envenoming causes a specific condition known as Capillary Leak Syndrome (CLS) [9]. Drowsiness, visual abnormalities, and respiratory failure are neurotoxicity signs [6]. Rarely, a snakebite can result in Guillain-Barre Syndrome (GBS) and Insulin Autoimmune Syndrome (IAS), which is treatable with plasmapheresis. For AKI, hemotoxic consequences, CLS, hemodialysis, and antisnake venom are used. But for many, plasmapheresis is a last-ditch option. Plasma exchange is an alternate method for removing venom toxins that are difficult to remove using dialysis or hemoperfusion. Venom toxins can be effectively removed from the blood by plasma exchange but not from other fluid compartments. Plasma exchange may help to reduce venom toxins in the blood compartment and may even cause venom toxins to be redistributed and removed from the extravascular region. Kornalik reported the first instance of plasma exchange being utilized to treat snake bite envenoming in 1990. Rasulov and Berdymuradov reported using plasma exchange in 1994 to treat three of 24 patients who had been bitten by snakes and become envenomed. Zengin et al. found plasmapheresis to be an effective intervention in a retrospective study of 37 snakebite patients with complications not resolving with antivenom administration [10]. Herein, we aim to conduct a systematic review to assess the effectiveness of plasmapheresis in snakebite cases and the situations under which it may be beneficial.

## Methods

### Search strategy

PROSPERO (ID: CRD42023459261) was used to register the study protocol. Prior to the start of the systematic review, a protocol was created using PRISMA (Preferred Reporting Items for Systematic Reviews and Meta-Analyses). A comprehensive search approach for articles from 1980 to July 2023 was carried out. PubMed, Embase via Elsevier, MEDLINE via Ovid, The Cochrane Library for Cochrane Reviews, Scopus, Web of Science, and Google Scholar were searched with the MeSH terms ‘bite’ or ‘snakebite’ or ‘envenomation’ or ‘snakebite envenoming’ and ‘plasmapheresis’ or ‘therapeutic plasma exchange.’ Additionally, we contacted the research authors for any missing data, manually checked the reference lists of the included studies, and utilised a database’s similar articles function.

### Study selection and screening

HP, NK, and JJ independently screened search results for eligibility. For articles eligible after screening, full texts were retrieved by HP, NK, and JJ, which HP, JJ, NK, AU, TS, AS and KS reviewed. HP and JJ screen the citation search. Discrepancies were resolved by consensus. The selection process was done as in Fig. 1 [11].

**Fig. 1:**
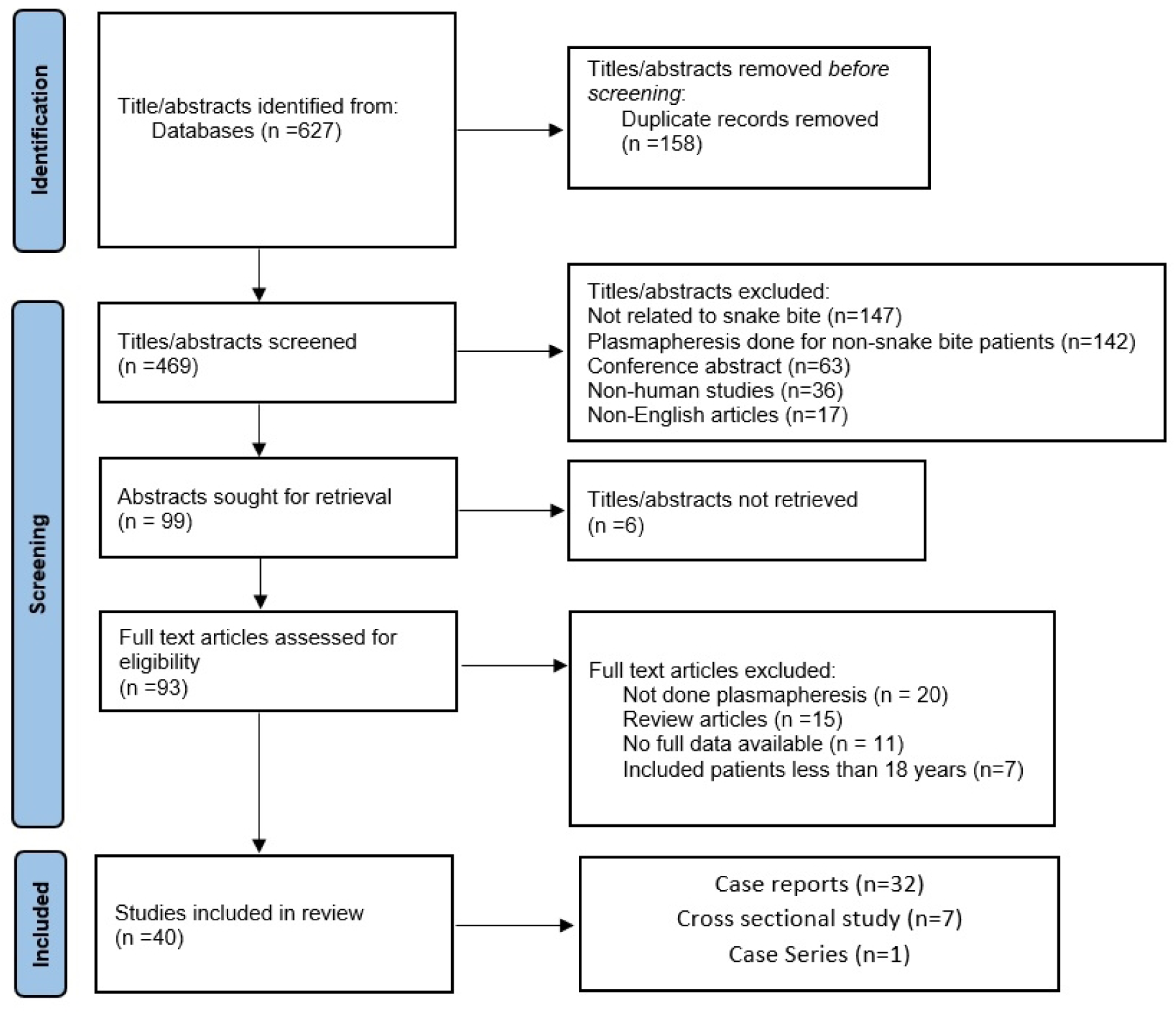
PRISMA above shows the flow of patients.

### Eligibility Criteria

This review aimed to find, assess, and synthesize all cross-sectional studies, case reports, or case series that used plasmapheresis for snakebite along with conventional Anti Snake Venom (ASV), atropine-neostigmine, calcium gluconate or hemodialysis. Only human cases aged more than 18 years, suspected or confirmed snakebites or those who fulfill indications of plasmapheresis were included in the review. We excluded Human cases under 18 years, studies reporting in vitro data only, review articles, or those with complete reporting overlap. The study was set in an Emergency Department or Intensive Care Unit as patients with complications mostly come here.

### Data extraction

HP and JJ were the authors who extracted the data. Each included study provided the following data regarding study characteristics and outcomes:

- Study types: observational studies
- The study’s methods included: Authors of the study, year, and study design
- The intervention(s) of interest: kind, frequency, and comparator(s) used: normal care
- Age, gender, and the outcomes listed above were characteristics of the participants’ studies.

### Assessment of the Risk of Bias

The risk of bias for each research was independently assessed by five review authors (NK, AN, TS, HP, JJ, AS and KS) using the Murad MH, Sultan S, Haffar S, Bazerbachi F. Methodological quality and synthesis of case series and case reports. BMJ Evid-Based Med. 2018;23(2): 60–3 [12].

### Outcome

The study’s primary outcomes were ‘improvement in renal dysfunction’ defined as patients becoming KDIGO 1 at discharge, ‘improvement in neurological dysfunction’ defined as Guillain Barre Syndrome disability score 0-2 at discharge, or ‘improvement in hematological dysfunction’ described as hemoglobin and platelets returning to age-appropriate normal limits without schistocytes following plasmapheresis done for snakebite. The secondary outcomes were discharge or death, morbidity (KDIGO 2 or3, Guillain Barre Syndrome disability score 3-5, no return to age-appropriate limits for hemoglobin and platelets), hospital stay duration, and number of plasmapheresis. This helps to determine the role of plasmapheresis and its effectiveness in the snakebite.

### Analysis

The mean and standard deviation for normal distributions, the median (interquartile range) for non-normal distributions of numerical variables, and percentages and proportions for categorical variables were the ways in which descriptive statistics were stated. The data was analysed using SPSS 23.0.

## Results

This systematic review includes 147 patients from 40 studies carried out between 1980 and July 2023. The population’s average age was 49.9 ±14 years, with 59.9% of patients being male.

### Main results

Regarding the types of snake bites, 77 cases involved unidentified snake species, while the hump-nosed viper (*Hypnale hypnale*, one of which is *Hypnale zara*) accounted for most known snakebites (42). This was followed by eight each for Russell’s viper (*Daboia russelii)* and saw-scaled viper (*five Echis carinatus,* one *Echis pyramidum, one Echis coloratus, one Echis sochureki)*, three pit vipers (*two Bothrops asper, one Crotalus cerastes)*, and other viper species (Proatheris superciliaris, *Cerastes cerastes*) accounting for two. Additionally, there were three krait bites (*one Bungarus caeruleus, one Bungarus multicinctus, one Bungarus candidus)*, one sea snake bite (*Hydrophis platurus)*, one Australian brown snake bite (*Pseudonaja nuchalis)*, one taipan bite (*Oxyuranus scutellatus*), and one Colubridae snake bite (*Notechis scutatus*).

A summary table showing study included, type of snake, indication for plasmapheresis and geographical location of snakebite are shown in Tabe 1.

**Table 1:** Summary Table.

| Sl.No | Author | Study | Snake | Indication | Geographical Location |
| --- | --- | --- | --- | --- | --- |
| 1. | Yildirim et al. [13] | Retrospective descriptive study | N/A | VICC | Turkey |
| 2. | Arora et al. [14] | Case report | *Daboia russelii* | TMA | India |
| 3. | Mohan et al. [15] | Case report | *Daboia russelii* | TMA | India |
| 4. | Zengin et al. [10] | Retrospective observational study | N/A | VICC | Turkey |
| 5. | Berber et al. [16] | Observational study | N/A | VICC | Turkey |
| 6. | Dineshkumar et al. [17] | Case report | *Daboia russelii* | TMA | India |
| 7. | Erkurt et al. [18] | Retrospective observational study | N/A | VICC | Turkey |
| 8. | Rathnayaka et al. [19] | Prospective observational study | *Hypnale hypnale* | TMA | Srilanka |
| 9. | Isbister et al. [20] | prospective observational study | *Pseudonaja nuchalis* | TMA | Australia |
| 10. | Rahmani et al. [21] | Case series | *Echis coloratus* | TMA | Israel |
| 11. | Obeidat et al. [22] | Case Report | *Echis carinatus* | TMA | Jordan |
| 12. | Keyler et al. [23] | Case Report | *Proatheris superciliaris* | TMA | USA |
| 13. | Kornalik et al. [24] | Case Report | *Bothrops asper* | VICC | Czechoslovakia. |
| 14. | Valenta et al. [25] | Case Report | *Echis pyramidum* | TMA | Czech Republic |
| 15. | Sampley et al. [26] | Case Report | N/A | DAH | India |
| 16. | Grace et al. [27] | Case Report | N/A | CLS | India |
| 17. | Hameed et al. [28] | Case Report | *Hydrophis platurus* | GBS | Pakistan |
| 18. | Cobcroft et al. [29] | Case Report | *Oxyuranus scutellatus* | TMA | Australia |
| 19. | Chuang et al. [30] | Case Report | *Bungarus multicinctus* | GBS | Taiwan |
| 20. | Akhilesh Kumar et al. [8] | Case Report | *Echis sochureki* | DAH | India |
| 21. | Withana et al. [31] | Case Report | *Hypnale hypnale* | TMA | Srilanka |
| 22. | Mitrakrishnan et al. [32] | Case Report | *Hypnale hypnale* | TMA | Srilanka |
| 23. | Rathnayaka et al. [33] | Case Series | *Daboia russelii* | TMA | Srilanka |
| 24. | Srivastava et al. [34] | Case Report | N/A | GBS | India |
| 25. | Reddy et al. [35] | Case Report | *Daboia russelii* | TMA | India |
| 26. | Cañas et al. [36] | Case Report | *Bothrops asper* | TMA | Colombia |
| 27. | Rathnayaka et al. [37] | Case report | Hypnale zara | TMA | Srilanka |
| 28. | Godavari et al. [38] | Case Report | N/A | TMA | India |
| 29. | Murthy et al. [39] | Case Report | N/A | TMA | India |
| 30. | Wijewickrama et al. [40] | Prospective observational study | *Hypnale hypnale* | TMA | Srilanka |
| 31. | Janarthanam et al. [41] | Case Report | *Crotalus cerastes* | GBS | India |
| 32. | Senthilkumaran et al. [42] | Case Report | *Bungarus caeruleus* | IAS | India |
| 33. | Priyankara et al. [43] | Case Report | *Daboia russelii* | TMA | Srilanka |
| 34. | Laothong et al. [44] | Case Series | *Bungarus candidus* | GBS | Thailand |
| 35. | Herath et al. [45] | case series | *Hypnale hypnale* | TMA | Srilanka |
| 36. | Bava et al. [46] | Case series | *Echis carinatus* | DAH, VICC, TMA | India |
| 37. | Elmoqaddem et al. [47] | Case report | *Cerastes cerastes* | VICC | Morocco |
| 38. | Casamento et al. [48] | Case Series | *Notechis scutatus* | TMA | Australia |

CLS, Capillary Leak Syndrome; DAH, Diffuse Alveolar Hemorrhage; GBS, Guillain-Barre Syndrome; IAS, Insulin Autoimmune Syndrome; TMA, Thrombotic Microangiopathy; USA, United States of America; VICC, venom-induced consumption coagulopathy

Among the indications for plasmapheresis, the majority (51%) was performed for VICC, while 42.2% of cases were for TMA and 3.4% for GBS. Three cases involved DAH, and one each involved CLS and IAS, all of which underwent plasmapheresis.

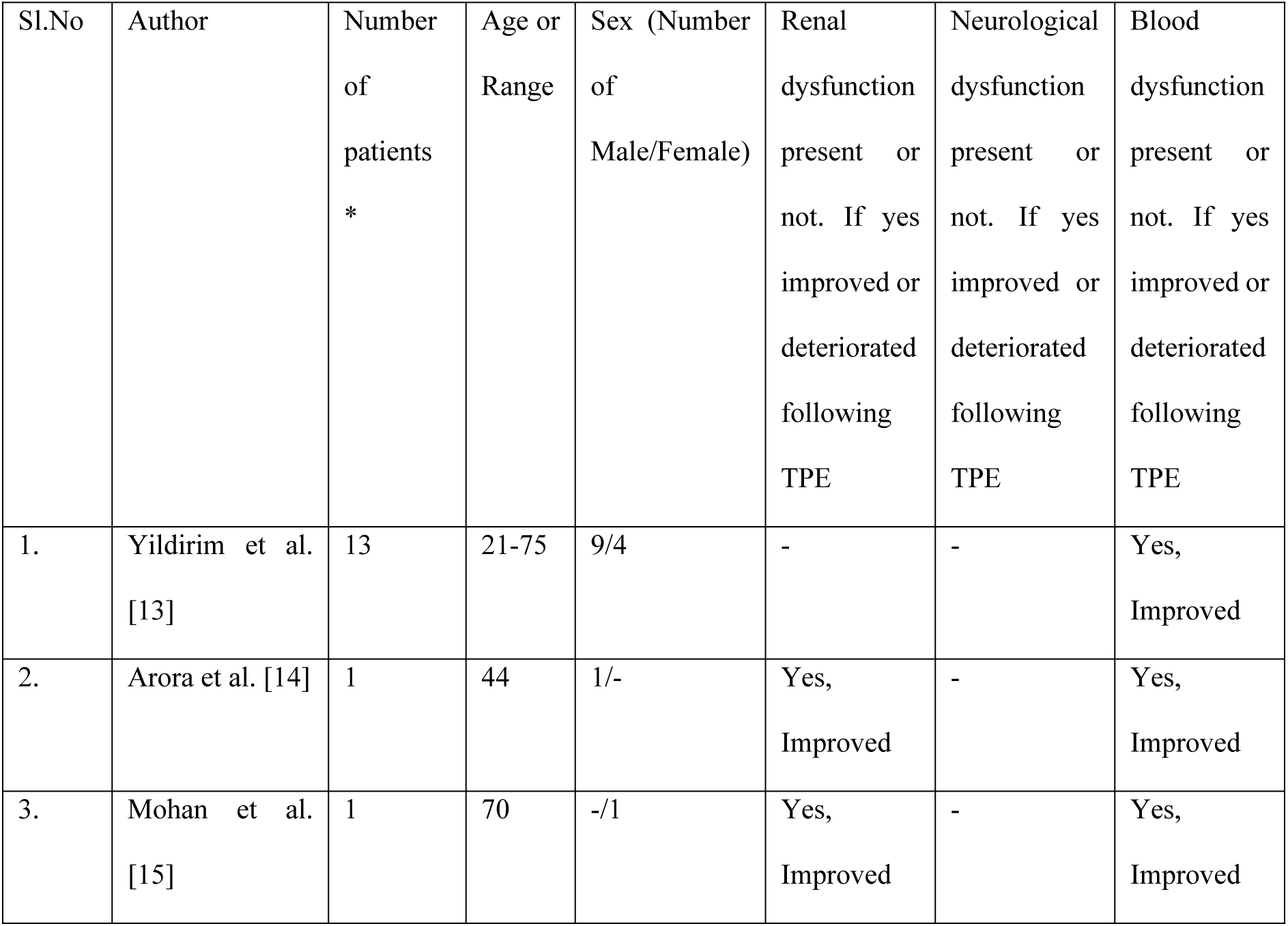
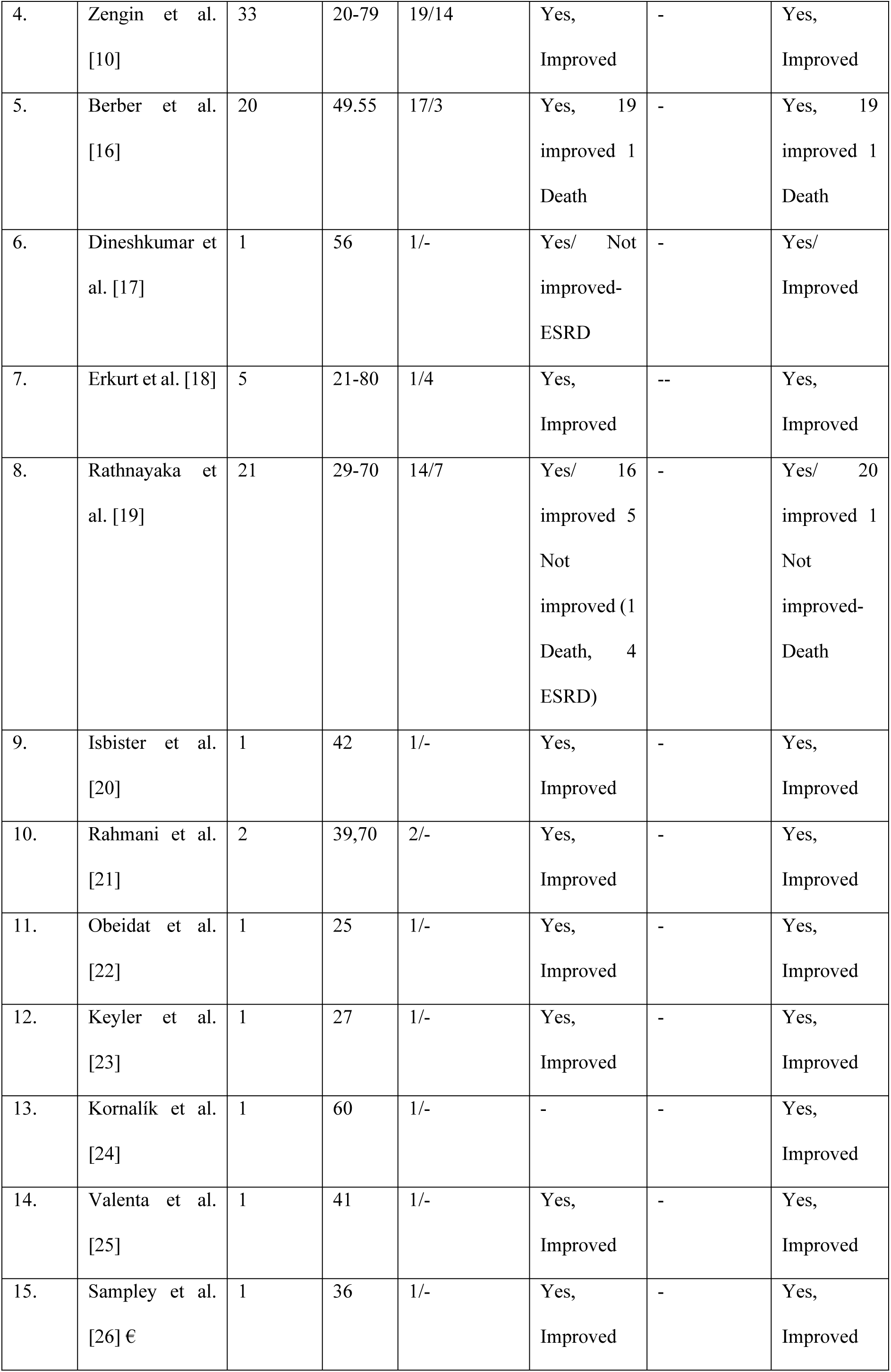
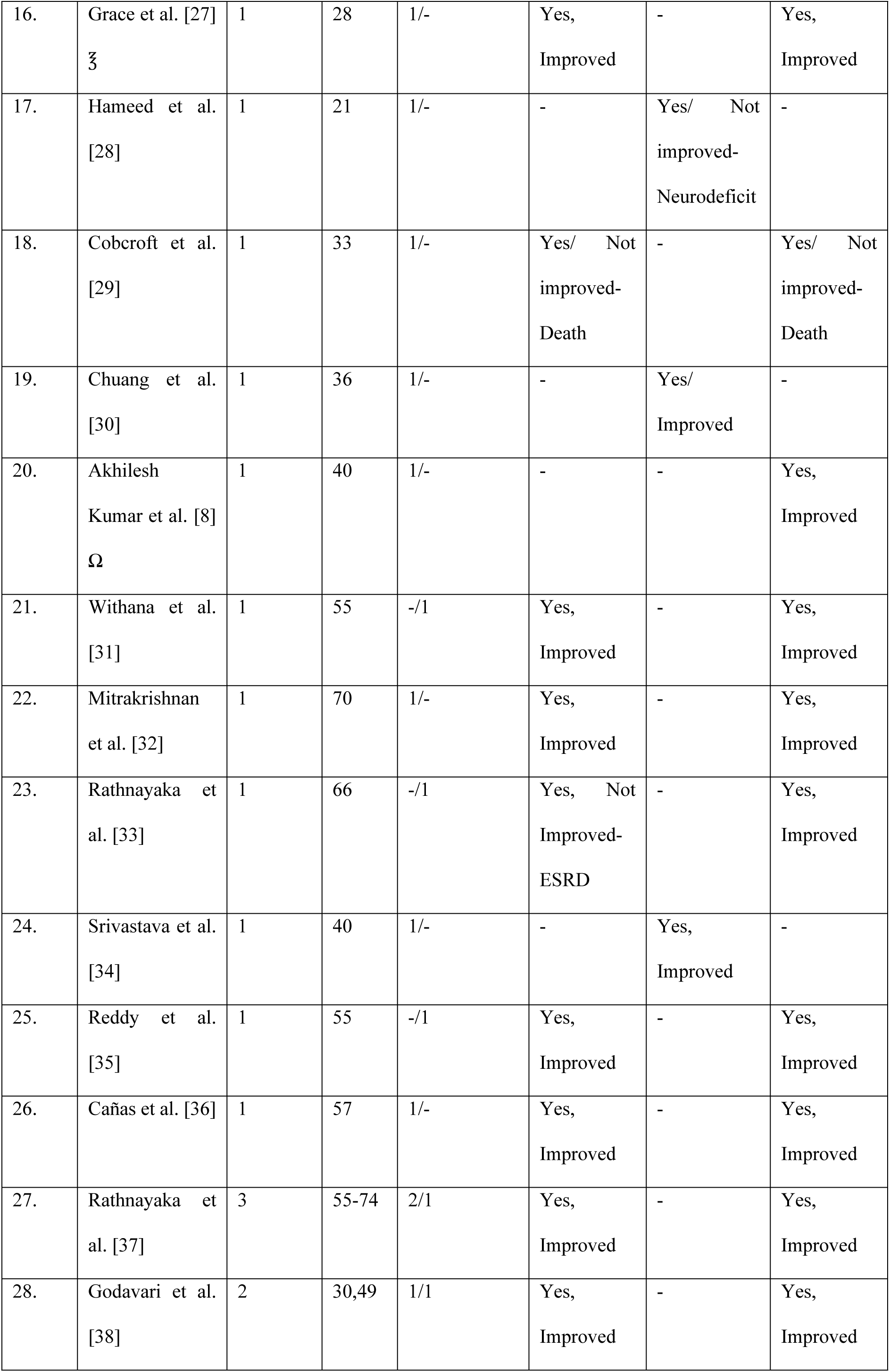
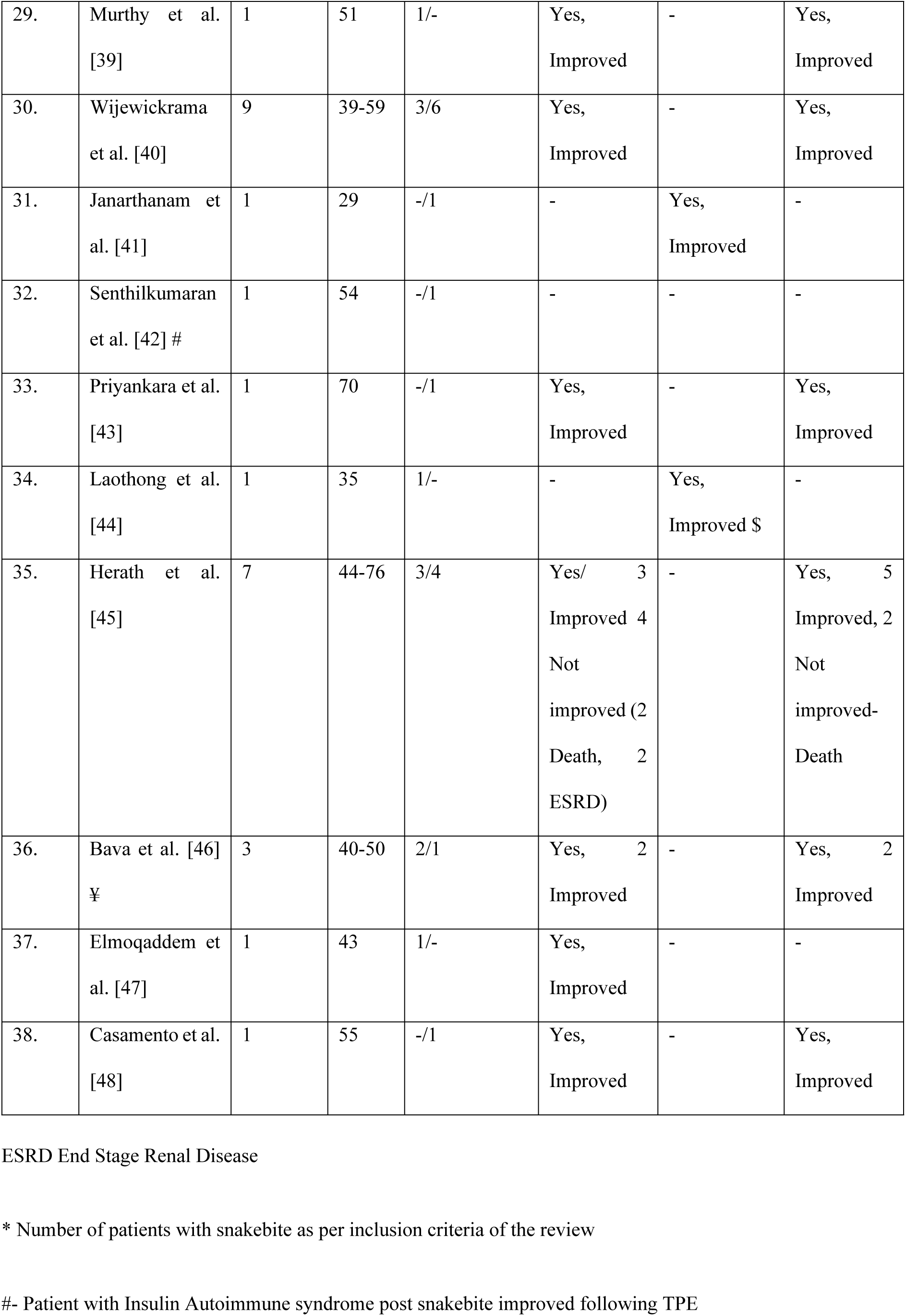
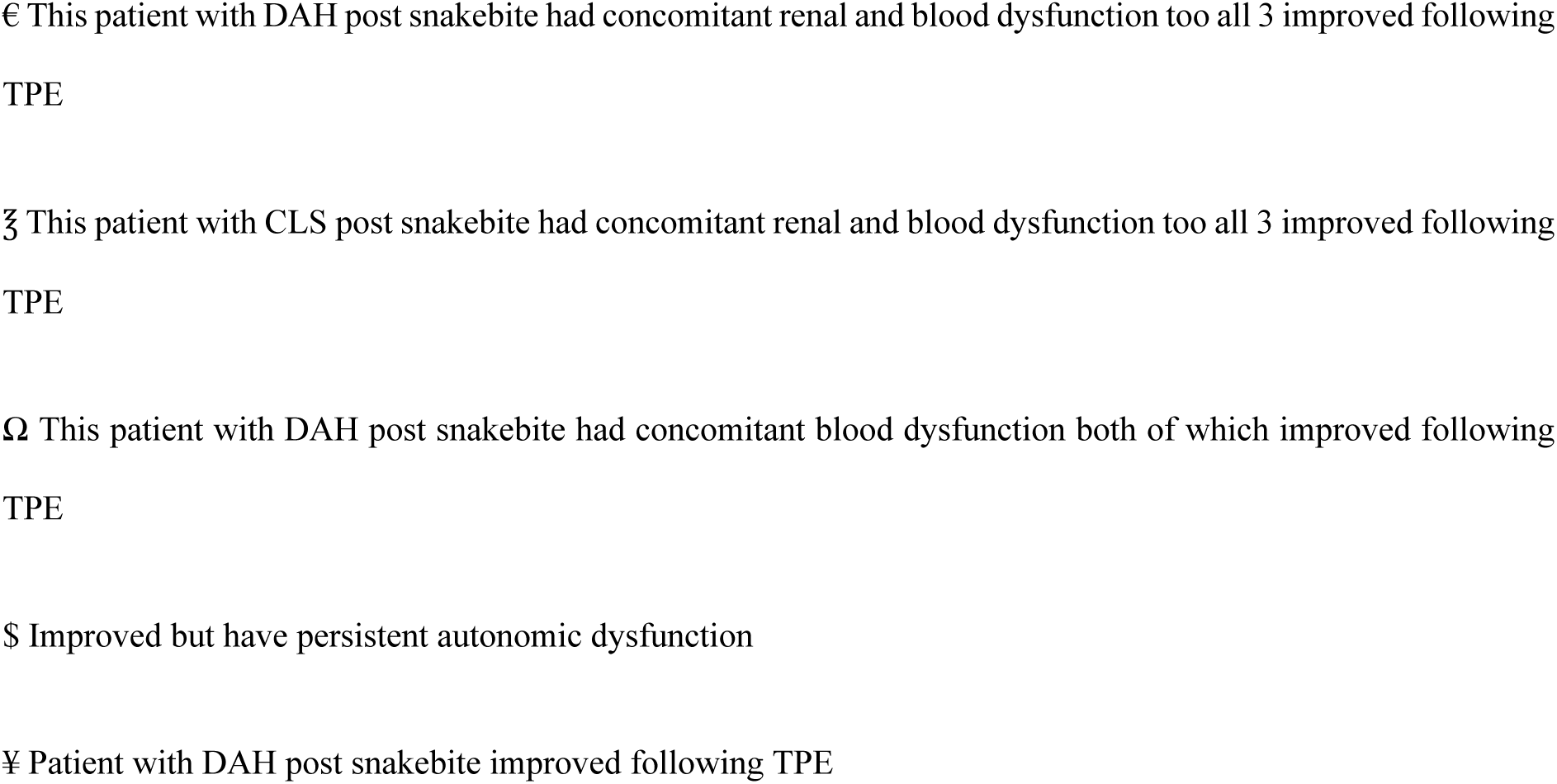

Of the 124 patients who developed renal dysfunction, 112 showed improvement after plasmapheresis. Five of the eight patients who developed neurological dysfunction showed improvement after plasmapheresis. Of the 136 patients who developed hematological derangements, 124 improved after plasmapheresis. Furthermore, one patient with IAS who developed autonomic dysfunction also improved after plasmapheresis.

From the available data, 90.5% of patients were discharged from the hospital, while nine patients succumbed to death. Among the patients, 86.4% did not develop residual clinical effects on discharge (no morbidity), 7.5% developed end-stage renal disease, two developed residual limb weakness, and one developed autoimmune dysfunction. Relation of indication for plasmapheresis and duration of hospital stay is shown in Fig. 2.

**Fig. 2:**
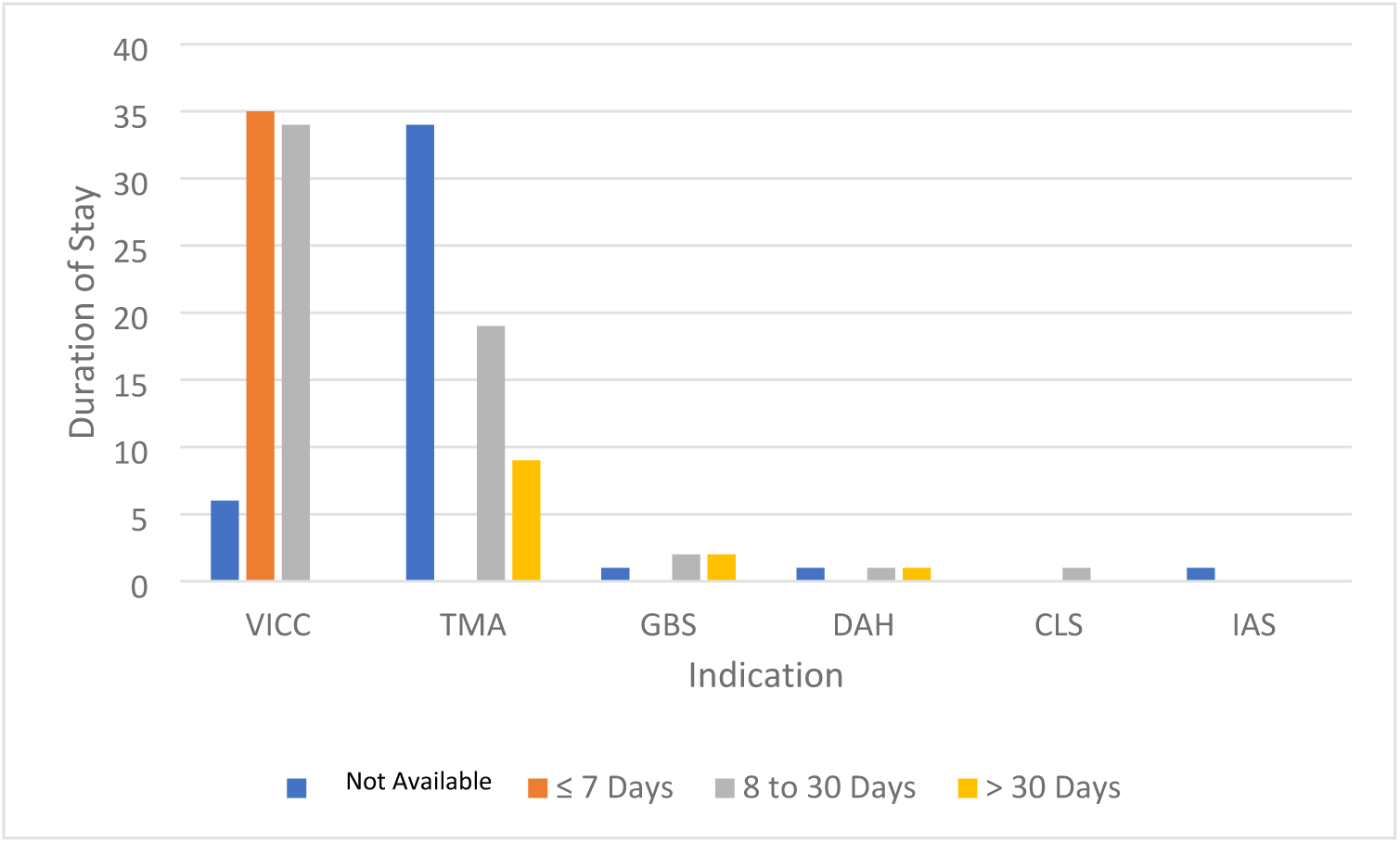
Bar diagram showing mean duration of hospital stay with plasmapheresis indications CLS, Capillary Leak Syndrome; DAH, Diffuse Alveolar Hemorrhage; GBS, Guillain-Barre Syndrome; IAS, Insulin Autoimmune Syndrome; TMA, Thrombotic Microangiopathy; VICC, venom-induced consumption coagulopathy

The duration of hospital stay showed high variability, ranging from 1 to 65 days, with a mean of 13.1 days. The number of plasmapheresis sessions ranged from 1 to 14, with a median of 2 and an interquartile range (IQR) of (1,4).

## Discussion

Snakebites are an occupational danger, with the majority of bites occurring on farms when working barefoot or wandering through the fields at night. The four families of venomous snakes are the Elapidae, Viperidae, Hydrophidae, and Colubridae. Elapid venom’s most prevalent clinical effects are neurotoxic, whereas viper venom causes vascular toxicity and nephropathy, and Hydrophiinae or sea snake venom is myotoxic [49].

AKI is a common complication of snakebite and a significant cause of death and morbidity. 75% of instances of AKI in this situation are caused by acute tubular necrosis (ATN), while 5–15% of cases are caused by acute interstitial nephritis (AIN) [38]. The prevalence of AKI following Russell’s viper (*Daboia russelii)* and *Echis carinatus* bites ranges from 13 to 32%. Pathogenesis of AKI in snake envenoming involves hemolysis, rhabdomyolysis, DIC, sepsis, hemodynamic abnormalities, and cell damage caused by the production of proinflammatory cytokines and vasoactive mediators [17,38].

### Venom induced consumption coagulopathy (VICC)

The hemotoxic venom activates coagulation pathways leading to VICC without thrombocytopenia and is characterized by severe bleeding without visible fibrin deposition, microvascular thrombosis, or end-organ failure, which results from snake toxins activating the coagulation pathway. This results in a prolonged prothrombin time, increased fibrinogen consumption, and elevated D-dimer levels, all overlapping with DIC. TMA appears in a subset of individuals with or without VICC whom a snake bite has envenoming. While the coagulopathy in VICC resolves rapidly, the triad of TMA (AKI, thrombocytopenia, and microangiopathic hemolytic anemia) persists for longer. Most cases of TMA following snake envenoming have been documented in Sri Lanka and Australia; very few cases have been reported in India. The proposed mechanism is that venom or its vascular endothelial toxins may function as factors related to VEGF or von Willebrand factor (vWF) activators and cause endothelium damage to initiate TMA. The role of ADAMTS-13, a VWF-cleaving protease, in snake bites requires more investigation [14,17].

The symptoms of TMA often begin 1-3 days after the bite and last longer than those of VICC, which typically resolves after 12–24 h of antivenom infusion. According to Bull et al., either the presence of intravascular fibrin or endothelial injury may predispose to fibrin deposition [50]. These data show that TMA may not always be the same as VICC. Certain species, including B. jararaca, induce TMA by up-regulating and activating the genes that produce inflammatory mediators such matrix metalloproteinase (MMP)-10, IL-8, VCAM-1, and E-selectin. Jararhagin, a hemorrhagic metalloproteinase discovered from B. jararaca venom, may cause a systemic proinflammatory response [51,52].

The ecarin enzyme converts prothrombin directly into meizothrombin and is primarily responsible for the pathophysiology of envenoming in members of the Echis genus. Disintegrins, including echistatin, ocellatin, and pyramidin, are other venom components that prevent platelet aggregation [53]. Hemorrhaging-like substances exacerbate prothrombotic activation and the production of fibrin deposits and lead to afibrinogenemia [25].

TPE for VICC was done in 51.1% of the study’s subjects, with 98.6% having their blood dysfunction improved.

### Thrombotic Micro Angiopathy (TMA)

TMA is a pathological diagnosis and is associated with acute kidney injury (AKI), Microangiopathic Hemolytic Anemia (MAHA) and thrombocytopenia. Thrombotic thrombocytopaenic purpura (TTP) and haemolytic uraemic syndrome (HUS) are the prototype diseases associated with TMA. The first is thrombotic thrombocytopenic purpura (TTP), which was discovered in 1925 in a 16-year-old female patient by Eli Moschcowitz. The second condition is hemolytic uremic syndrome (HUS), which was discovered by Gasser in 1955 in five newborns who had renal failure, thrombocytopenia, and hemolytic anaemia as a result of a diarrheal sickness [54,55]. Bite-related HUS or HUS-like alterations in Western Australia have been linked to gwadar (*Demansia nuchalis nuchalis*), dugite (*Demansia nuchalis affinis*), and Russell’s viper (*Daboia russelii)* bites in India. The difference between HUS and TTP is a little arbitrary because there is much overlap between the two, and it would be preferable to think of them as a continuum of a related disease process [29].

TMA is not included in the categories of complement factor dysregulation or ADAMTS 13 deficiency. The primary cause of TMA pathogenesis, according to the study, is endothelial cell damage or dysfunction, which exposes thrombogenic components to coagulation factors and platelets. Endothelial damage occurs as a result of uncontrolled inflammation and/or genetic or acquired abnormalities of complement or hemostasis mediators, as shown in TMA. TPE may be considered as a therapy option for such people if TMA is shown to be the consequence of snake envenoming [15]. TPE for TMA was done in 42.2 % of the study’s subjects, with 77.76 % having their renal dysfunction improved and 92.5% have their blood dysfunction improved.

### Guillain Barre Syndrome (GBS)

The first instance of GBS associated with a snakebite was reported by Chuang et al. in 1996. Four weeks after being bitten by a Bungarus multicinctus, sensorimotor axonal-type polyneuropathy was identified by EMG/NCS. After therapy (plasmapheresis and 500 mg/day of methylprednisolone for five days), the patient’s condition improved [30]. In 2010, Srivastava et al. reported that EMG/NCS revealed motor and sensory neuropathy five weeks after the bite, indicating demyelination with subsequent axonal degradation. Plasmapheresis aided the patient’s recovery [34]. Neil et al. (2011) described a patient who experienced paraesthesia and quadriparesis 12 days after being bitten by a *Vipera aspis* snake. The researchers discovered molecular similarity between venom proteins and neuronal GM2 gangliosides, hypothesising an immunological explanation rather than direct venom toxicity for this relationship [28,56]. TPE for GBS was performed on 3.4% of the study’s individuals, with four of them having neurological dysfunction improved and one having neurodeficit. One of those who improved had persisting autonomic dysfunction.

### Diffuse Alveolar Hemorrhage (DAH)

DAH is a potentially fatal pulmonary complication in various conditions characterized by hypoxemia caused by alveolar infiltrates, which leads to alveolar hemorrhage and necessitates rapid treatment and lifesaving measures. DAH’s causes can be immunological and non-immune. The pathophysiology includes widespread alveolar destruction, alveolar hemorrhage, and antibody-mediated pulmonary capillaritis. Direct venom action and/or an insult mediated by an antibody might account for the delayed DAH by causing widespread alveolar injury. Sampley et al. described DAH in viper envenoming during the first week, with a normal coagulation profile controlled with methylprednisolone and plasmapheresis. The effective use of plasmapheresis and methylprednisolone in DAH on day three following a hump-nosed pit viper (*Hypnale hypnale)* bite, including VICC and thrombotic microangiopathy, was described by Srirangan et al. Therefore, plasma exchange has a limited role in the treatment of snakebite despite evidence supporting its use in thrombotic microangiopathy [8,26]. TPE was performed on three individuals with DAH, two of whom had concurrent blood malfunction and one of whom had concurrent renal impairment. All of which improved after TPE.

### Capillary Leak Syndrome (CLS)

Dr. Bayard Clarkson discovered CLS, also known as Clarkson’s illness, in 1960. Its defining features are hemoconcentration, hypotension, generalized edema, and hypoalbuminemia without albuminuria [57]. Capillary hyperpermeability and plasma extravasation are the essential mechanisms. CLS can develop as a primary (idiopathic) disorder or a complication of other illnesses. Snake venom contains toxins and enzymes that promote capillary permeability and cause plasma to escape from the intravascular region. The clinical appearance of CLS goes through three stages. First, a prodromal phase of irritation, myalgia, thirst, and syncope. Second, the capillary leakage phase is characterized by intravascular fluid extravasation. Third, complication phase-renal failure is a frequent complication [27]. There are no recognized diagnostic criteria, but with minor revisions, the Thomas and Kumar criteria can be used for diagnosis.

Treatment includes theophylline, leukotriene antagonists, terbutaline, plasmapheresis, corticosteroids, and intravenous immunoglobulin (IG), disease-modifying therapy medications. The outlook for CLS is still quite bad because there is no established treatment. We hypothesized that the patient’s favorable result may have been influenced by early CLS detection, timely plasmapheresis implementation, and the use of fresh frozen plasma [58]. TPE was done for one patient with CLS post snakebite who had concomitant renal and blood dysfunction too all of which improved following treatment.

### Insulin Autoimmune Syndrome (IAS)

In the early 1970s, Hirata and colleagues described Insulin Autoimmune Syndrome (IAS). Postprandial hypoglycemia, increased insulin levels, and detectable insulin antibodies are all indications of IAS. These antibodies, most often IgG, are considered to bind to circulating insulin and inhibit its interaction with its receptor. There have been a few accounts of a snakebite-inducing IAS. Many biomolecules in snake venom have the potential to be antigenic. Common krait’s (*Bungarus caeruleus)* venom may contain a compound with structural resemblance to endogenous insulin, leading to the production of antibodies against its epitope, or venoms could include substances that reduce disulfide and cause the production of monomers. The existence of the HLA-DRB1*0406 allele has been linked to the onset of IAS. Dextrose infusions, corticosteroids, somatostatin, diazoxide, azathioprine, rituximab, metformin, dietary changes, and plasmapheresis have all been used to treat IAS. Plasmapheresis rapidly decreased the patient’s autoantibody level and reversed their condition. Therefore, once the diagnosis has been made, particularly for individuals with krait (*Bungarus caeruleus)* envenoming, early usage of plasmapheresis will be beneficial [42]. One patient with Insulin Autoimmune syndrome post snakebite was there in study who improved following TPE

### Plasmapheresis

Plasma exchange is an alternate method for removing venom toxins that are difficult to remove using dialysis or hemoperfusion. Venom toxins can be effectively removed from the blood by plasma exchange but not from other fluid compartments. Plasma exchange may help to reduce venom toxins in the blood compartment and may even cause venom toxins to be redistributed and removed from the extravascular region. Kornalik reported the first instance of plasma exchange being utilized to treat snake bite envenoming in 1990. Rasulov and Berdymuradov reported using plasma exchange in 1994 to treat three of 24 patients who had been bitten by snakes and become envenomed. Zengin et al. found plasmapheresis to be an effective intervention in a retrospective study of 37 snakebite patients with complications not resolving with antivenom administration [10].

Plasmapheresis has been utilized to treat TMA caused by snake bites in the absence of antivenom. Plasma exchange therapy should be started before a laboratory test can definitively establish the diagnosis because of the substantial mortality risk associated with TTP. This lack of supporting evidence shows that plasma exchange has only a minor role in snakebite envenoming. Therapeutic plasma exchange for snakebite envenoming is classified as Category III, grade 2 C by the American Society for Apheresis (ASFA) (poor recommendation, low-quality or extremely low-quality evidence) [8].

According to the ASFA criteria, acquired TTP is a category I indication for TPE (grade 1A evidence). TPE has two advantages: it eliminates autoantibodies against ADAMTS13 and infuses donor plasma with exogenous ADAMTS13 enzyme. The ASFA classified (Shiga like Toxin-Producing E.coli) STEC-HUS as category IV. The argument for using TPE in aHUS is that when allogeneic donor plasma is utilized as the replacement fluid for the exchange; this treatment will eliminate the faulty complement proteins and autoantibodies against complement factor H (CFH). According to ASFA recommendations, TPE is a second-line treatment for patients with additional complement mutations (category II, grade 2C) and a first-line treatment for those who have anti-CFH autoantibodies (category I, grade 2C) [23,59].

Other indications for plasmapheresis include Guillain-Barré syndrome, ANCA-associated and Goodpasture glomerulonephritis, chronic demyelinating polyradiculoneuropathy, recurrent focal segmental glomerulosclerosis, hyperviscosity in monoclonal gammopathies, myasthenia gravis, and encephalitis (Category 1). Acute encephalomyelitis, cardiac and stem cell transplantation desensitisation, antiphospholipid syndrome, cryoglobulinemia, cardiomyopathy, Hashimoto encephalopathy, Lambert-Eaton syndrome, multiple sclerosis, and myeloma nephropathy (Category 2) are also included. Category 3 indications include acute liver failure, autoimmune hemolytic anaemia, burn shock, newborn lupus, chronic encephalitis, heparin-induced thrombocytopenia, pancreatitis, refractory thrombocytopenia, paraneoplastic syndromes, sepsis, and vasculitis. Systemic amyloidosis, dermatomyositis/polymyositis, and lupus nephritis are all classified under Category 4 [60].

Contraindications for therapeutic plasmapheresis include lack of central line access or large bore peripheral lines, hemodynamic instability or septicemia, a known allergy to fresh frozen plasma or replacement colloid/albumin, or heparin; relative contraindications include hypocalcemia and angiotensin-converting enzyme (ACE) inhibitor use within the previous 24 hours [61]. Complications of plasmapheresis are hypocalcemia or hypomagnesemia due to citrate anticoagulation, hypotension, flushing, nausea and vomitinghypothermia, fluid and electrolyte imbalances, transfusion reactions, thrombocytopenia and hypofibrinogenemia causing bleeding diatheses [62]. Plasmapheresis is an effective treatment for a variety of acute and chronic conditions. It may also be used to help prepare for operations and surgeries, as well as to support postoperative recovery and minimise respiration periods. In general, the risks and side effects are minor, with the most prevalent being brief hypotension and associated symptoms [63].

However, antivenom should be administered as soon as feasible, and plasma exchange should be considered as a supplemental therapy strategy for patients with severe envenoming, particularly in managing hematologic issues and limb preservation/salvage techniques. We believe that plasma exchange treatment is advantageous because it eliminates toxins and inflammatory mediators. If the envenoming from a snake bite cannot be adequately treated using standard methods, plasma exchange may be explored as part of the treatment [10].

So, we propose that plasmapheresis could be used in snakebites in complications such as VICC, TMA, GBS, DAH, CLS, and IAS. Given the rarity of presentation and lack of evidence, plasma exchange could be a lifesaving intervention in some circumstances. The effectiveness of plasma exchange in treating snake bite envenoming should be clarified by conducting extensive randomized controlled trials.

### Limitations and strengths

We have included only case series, case reports, and cross-sectional studies as it is challenging to carry out a randomized controlled trial, case-control, or cohort study. The risk of bias was assessed by a nonconventional method using Murad et al. Comparative studies of hemodialysis versus plasmapheresis are required. Plasmapheresis is a costly procedure, and widespread availability is a concern.

The study’s strength is that it is one of the first to analyze the role of plasmapheresis in snake bite and to list the indications when plasmapheresis may be used in snakebites.

## Conclusion

Snakebites are serious occupational hazards particularly affecting people in agricultural contexts. Various snake venoms have diverse toxicological profiles, including neurotoxicity, vascular toxicity, nephropathy, and myotoxicity. Acute kidney damage (AKI) is a common and serious complication of snakebite, usually caused by acute tubular necrosis. Hemotoxic venoms can cause coagulopathies, resulting in thrombotic microangiopathy (TMA), which is characterised by acute kidney injury, thrombocytopenia, and microangiopathic hemolytic anaemia. Plasma exchange (plasmapheresis) has emerged as a potential therapeutic option for snakebite complications such as TMA, Guillain-Barré syndrome, diffuse alveolar haemorrhage (DAH), and capillary leak syndrome (CLS). Despite limited evidence, plasmapheresis can be life-saving by removing toxins and inflammatory mediators from the circulation. It is especially useful when conventional therapies are insufficient. Randomised controlled trials are required for establishing the effectiveness of plasmapheresis in snakebite envenoming, however its use as an adjuvant treatment shows promise for improving patient outcomes in severe envenoming.

## Data Availability

All data underlying are provided in attachments as supporting information without restriction

## Acknowledgements

None

## Author Contributions

NK had full access to all of the study’s data and accepts responsibility for the data’s integrity and the correctness of the data analysis. As co-first authors, HP and JRJ contributed equally.

Corresponding author: NK

Concept and design: HP, NK, JRJ, AU, TS, AS, KS.

Acquisition, analysis, or interpretation of data: HP, JRJ, AU.

Drafting of the manuscript: HP, NK, JRJ, AS, AU, TS, KS

Critical revision of the manuscript for important intellectual content: HP, JRJ, NK, AS, KS, AU, TS

Statistical analysis: AU, HP, JRJ.

Administrative, technical, or material support: NK.

Supervision: HP, JRJ, NK, AS, KS, TS, AU

## Availability of data and materials

The datasets generated during and/or analysed during the current study are available from the corresponding author on reasonable request.

## Declarations

Ethics approval and consent to participate

Not applicable.

## Consent for publication

Not applicable

## Author Information

HP, Junior Resident, Department of Emergency Medicine, All India Institute of Medical Sciences, Rishikesh, India,

NK, Head of Department, Department of Emergency Medicine, All India Institute of Medical Sciences, Rishikesh, India,

JRJ, Junior Resident, Department of Emergency Medicine, All India Institute of Medical Sciences, Rishikesh, India,

AU, Junior Resident, Department of Community and Family Medicine, All India Institute of Medical Sciences, Rishikesh, India,

TS, Senior Resident, Department of Emergency Medicine, All India Institute of Medical Sciences, Rishikesh, India,

AS, Junior Resident, Department of Paediatrics, All India Institute of Medical Sciences, Rishikesh, India,

KS, MD, Senior Resident, Department of Emergency Medicine, All India Institute of Medical Sciences, Rishikesh, India,

